# EpiLPS: a fast and flexible Bayesian tool for near real-time estimation of the time-varying reproduction number

**DOI:** 10.1101/2021.12.02.21267189

**Authors:** Oswaldo Gressani, Jacco Wallinga, Christian Althaus, Niel Hens, Christel Faes

**Affiliations:** Interuniversity Institute for Biostatistics and statistical Bioinformatics (I-BioStat), Data Science Institute, Hasselt University, Belgium; Centre for Infectious Disease Control, National Institute for Public Health and the Environment, Bilthoven, The Netherlands; Department of Biomedical Data Sciences, Leiden University Medical Centre, Leiden, The Netherlands; Institute of Social and Preventive Medicine, University of Bern, Bern, Switzerland; Centre for Health Economics Research and Modelling Infectious Diseases, Vaxinfectio, University of Antwerp, Belgium

## Abstract

In infectious disease epidemiology, the instantaneous reproduction number *R*(*t*) is a timevarying metric defined as the average number of secondary infections generated by individuals who are infectious at time *t*. It is therefore a crucial epidemiological parameter that assists public health decision makers in the management of an epidemic. We present a new Bayesian tool for robust estimation of the time-varying reproduction number. The proposed methodology smooths the epidemic curve and allows to obtain (approximate) point estimates and credible envelopes of *R*(*t*) by employing the renewal equation, using Bayesian P-splines coupled with Laplace approximations of the conditional posterior of the spline vector. Two alternative approaches for inference are presented: (1) an approach based on a maximum a posteriori argument for the model hyperparameters, delivering estimates of *R*(*t*) in only a few seconds; and (2) an approach based on a MCMC scheme with underlying Langevin dynamics for efficient sampling of the posterior target distribution. Case counts per unit of time are assumed to follow a Negative Binomial distribution to account for potential excess variability in the data that would not be captured by a classic Poisson model. Furthermore, after smoothing the epidemic curve, a “plug-in” estimate of the reproduction number can be obtained from the renewal equation yielding a closed form expression of *R*(*t*) as a function of the spline parameters. The approach is extremely fast and free of arbitrary smoothing assumptions. EpiLPS is applied on data of SARS-CoV-1 in Hong-Kong (2003), influenza A H1N1 (2009) in the USA and current SARS-CoV-2 pandemic (2020-2021) for Belgium, Portugal, Denmark and France.

**Author summary:** The instantaneous reproduction number *R*(*t*) is a key metric that provides important insights into an epidemic outbreak. We present a flexible Bayesian approach called EpiLPS (Epidemiological modeling with Laplacian-P-splines) for smooth estimation of the epidemic curve and *R*(*t*). Computational speed and absence of arbitrary assumptions on smoothing makes EpiLPS an interesting tool for near real-time estimation of the reproduction number. An R software package is available (https://github.com/oswaldogressani).

## 1 Motivation

The instantaneous reproduction number *R*(*t*) is a time-varying metric defined as the average number of secondary cases generated by infectious individuals at time *t*. During epidemic outbreaks, *R*(*t*) provides a snapshot (often on a daily basis) that quantifies the extent to which a given infectious disease is transmissible in a population and is therefore an important tool that assists governmental organizations in the management of a public health crisis. The reproduction number is also a good proxy for measuring the real-time growth phase of an epidemic and as such, constitutes a key signal about the severity of the outbreak and the required control effort. For this reason, having a robust, accurate and timely estimator of *R*(*t*) is a crucial matter that has attracted considerable interest in developing new statistical approaches during the last two decades as summarized in White et al. (2021). The paper of Gostic et al. (2020) compares several methods for estimating *R*(*t*) and gives clear insights about the main challenges and obstacles that have to be faced. They recommend the method of Cori et al. (2013) and its associated EpiEstim package (Cori, 2021) as an appropriate and accurate tool for near real-time estimation of the instantaneous reproduction number. Another recent approach is proposed in Parag (2021), where a recursive Bayesian smoother based on Kalman filtering is used to derive a robust estimate of *R*(*t*) in periods of low incidence. The EpiNow2 package (Abbott et al., 2020) also provides interesting extensions and implementations of current best practices for precise estimation and forecast of the reproduction number using a Bayesian latent variable framework. Spline based approaches have shown to be an interesting tool for flexible modeling of the reproduction number. Azmon et al. (2014) use penalized radial splines for estimating *R*(*t*) under a Bayesian setting with misreported data and Gressani et al. (2021) accelerated the computational implementation by replacing the MCMC scheme with Laplace approximations. From a frequentist perspective, Pircalabelu (2021) uses truncated polynomials and radial basis splines to model the series of new infections and a derivative thereof as a candidate estimator for the reproduction number.

In this article, we propose a new Bayesian approach termed “EpiLPS” for estimating *R*(*t*) based on case incidence data and the serial interval distribution (the time elapsed between the onset of symptoms in an infector and the onset of symptoms in an infected case). Our estimator of *R*(*t*) is based on epidemic renewal-equations (Fraser, 2007; Wallinga and Lipsitch, 2007) and Laplacian-P-splines (LPS) smoothing of the mean number of new cases by day of reporting. Time series of new cases by day of reporting are assumed to follow a Negative Binomial distribution to account for potential excess variability in the data that would not be captured by a classic Poisson model. Algorithms related to Laplace approximations and evaluations of B-spline bases are coded in C++ and embedded in the R language through the Rcpp package (Eddelbuettel et al., 2011), making computational speed another key strength of EpiLPS as *R*(*t*) can be estimated in seconds. In addition, EpiLPS can also be used to obtain a smoothed estimate of the epidemic curve that can be of potential interest to further visualize an epidemic outbreak.

The proposed Bayesian methodology is based on a latent Gaussian model for the B-spline amplitudes and opens up two possible paths for inference. The first is called LPSMAP, a fully “sampling free” approach based on Laplace approximations to the conditional posterior of B-spline coefficients. The hyperparameter vector is fixed at its *maximum a posteriori* and credible envelopes of *R*(*t*) are computed via the “delta” method. The second path is called LPSMALA and is an MCMC-based approach with Langevin diffusions for efficient exploration of the posterior distribution of latent variables. The latter approach is computationally heavier than LPSMAP but has the merit of taking into account the uncertainty surrounding the hyperparameters. The underlying Metropolis-within-Gibbs structure keeps the practical implementation to a fairly simple level and the computational cost is reasonable even for long chain lengths.

Compared to existing methods, EpiLPS resembles EpiEstim from a methodological point of view in the sense that *R*(*t*) is estimated from incidence time series and a serial interval distribution, yet the two approaches fundamentally differ in many aspects. First, the methodology of Cori et al. (2013) assumes that incidence at time *t* is Poisson distributed, while EpiLPS assumes a Negative Binomial model. Second, as our approach uses penalized spline based approximations, the prior specifications are imposed on the roughness penalty parameter and not directly on *R*(*t*) as in EpiEstim. Third and most importantly, EpiLPS is free of any sliding window specification. An R package for EpiLPS has been developed and is available at https://github.com/oswaldogressani. The software also allows to compute the Cori et al. (2013) estimate of *R*(*t*) for the sake of comparison.

The manuscript is organized as follows. Section 2 aims at presenting the Laplacian-P-splines model for smoothing count data. We show how the Laplace approximation applies to the conditional posterior of the B-spline amplitudes and also derive the (approximate) posterior of the hyperparameter vector to be optimized. This yields the *maximum a posteriori* estimate of the spline vector via Laplacian-P-splines (LPSMAP). Section 3 uses LPSMAP and proposes a “plug-in” estimate of *R*(*t*) based on renewal equations. Approximate credible intervals for *R*(*t*) will also be shown in this section. Section 4 shows an alternative path for estimation of *R*(*t*) based on Markov chain Monte Carlo (MCMC). The latter approach uses Langevin dynamics for efficient sampling of the target posterior distribution. This approach is termed LPSMALA for “Laplacian-P-splines with a Metropolis-adjusted Langevin algorithm”. Section 5 is devoted to assess the performance of EpiLPS in various simulation scenarios and make comparisons with EpiEstim. In Section 6, we apply EpiLPS to real world epidemic outbreaks. Finally, Section 7 concludes with a discussion.

## 2 Methodology behind EpiLPS

### 2.1 Negative Binomial model for case incidence data

Let {*y*_*t*_, *t* = 1, …, *T*} be a time series of counts during an epidemic of *T* days with *y*_*t*_ ∈ ℕ (set of non-negative integers) denoting the total number of new contaminations on day *t*. We assume that the number of cases on day *t* follows a Negative Binomial distribution *y*_*t*_ ∼ NegBin(*μ*(*t*), *ρ*), with 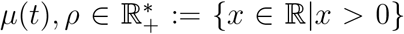 and probability mass function (see e.g. Anscombe, 1950; Piegorsch, 1990):

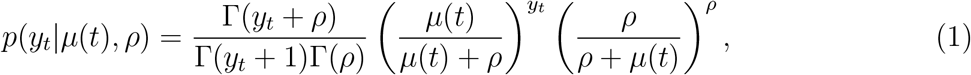

where Γ(·) is the gamma function. Under the above parameterization, we have 𝔼(*y*_*t*_) = *μ*(*t*) and 𝕍(*y*_*t*_) = *μ*(*t*) + *μ*(*t*)^2^*/ρ*, so that 1*/ρ* is the parameter responsible for overdispersion (variance larger than the mean) that is absent in a Poisson setting. In the limiting case lim_*ρ→*+*∞*_ 𝕍(*y*_*t*_) = *μ*(*t*) = 𝔼(*y*_*t*_) and we recover the Poisson model. We assume that *μ*(*t*) evolves smoothly over the time course of the epidemic and model it with cubic B-splines (Eilers and Marx, 1996):

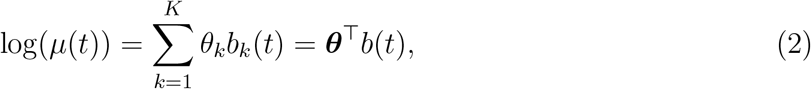

where ***θ*** = (*θ*_1_, …, *θ*_*K*_)^*⊤*^ is the vector of B-spline amplitudes to be estimated and *b*(·) = (*b*_1_(·), …, *b*_*K*_(·))^*⊤*^ is a cubic B-spline basis defined on the domain *𝒯* = [*r*_*l*_, *T*], where *r*_*l*_ is a lower bound on the time axis, typically the first day of the epidemic (i.e. *r*_*l*_ = 1). The philosophy behind P-splines consists in specifying a “large” number *K* of basis functions together with a discrete roughness penalty λ***θ***^*⊤*^*P****θ*** as a counterforce to the induced flexibility of the fit. The parameter *λ >* 0 acts as a tuning parameter calibrating the “degree” of smoothness and 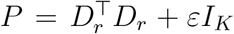 is a penalty matrix built from *r*th order difference matrices *D*_*r*_ of dimension (*K − r*) *× K* perturbed by an *ε*-multiple (here *ε* = 10^−6^) of the *K*-dimensional identity matrix *I*_*K*_ to ensure full rankedness. The reader is redirected to Eilers and Marx (2021) for a complete textbook treatment of P-splines. Following Lang and Brezger (2004), we impose a Gaussian prior on the spline vector 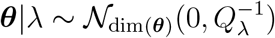, with precision matrix *Q*_*λ*_ = *λP*. For full Bayesian inference, the following priors are imposed on the model hyperparameters. Following Jullion and Lambert (2007), a robust Gamma prior is specified for the roughness penalty parameter *λ*|*δ* ∼ 𝒢 (*ϕ/*2, (*ϕδ*)*/*2), where 𝒢(*a, b*) is a Gamma distribution with mean *a/b* and variance *a/b*^2^, *ϕ* = 2 and *δ* is an additional dispersion parameter with hyperprior *δ* ∼ 𝒢(*a*_*δ*_ = 10, *b*_*δ*_ = 10). Finally, the following uninformative prior is imposed on the overdispersion parameter *ρ* ∼ 𝒢(*a*_*ρ*_ = 0.0001, *b*_*ρ*_ = 0.0001). Let ***η*** := (*λ, ρ*)^*⊤*^ denote the vector of hyperparameters. The full Bayesian model is thus:

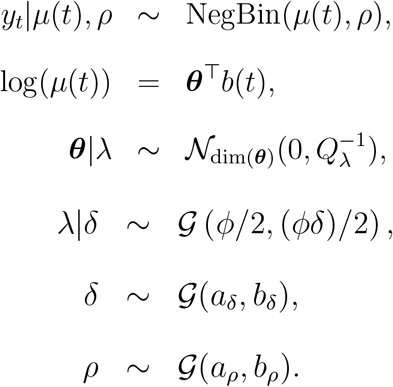

### 2.2 Laplace approximation to the conditional posterior of *θ*

The log-likelihood for the Negative Binomial model is given by:

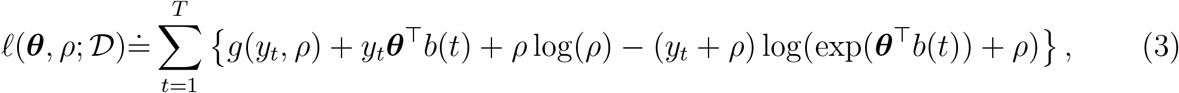

with *g*(*y*_*t*_, *ρ*) = log Γ(*y*_*t*_ + *ρ*) *−* log Γ(*ρ*) and ≐ denoting equality up to an additive constant. The gradient of the log-likelihood with respect to the spline coefficients is:

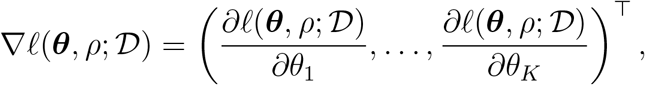

where:

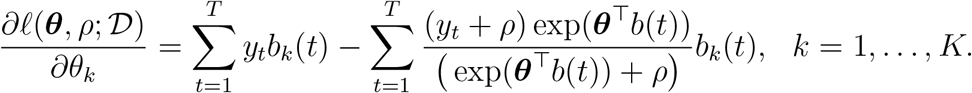

The Hessian of the log-likelihood with respect to the B-spline amplitudes is:

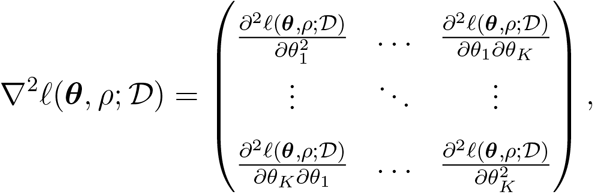

with entries:

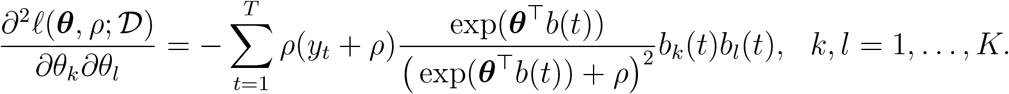

Using Bayes’ rule, the conditional posterior of ***θ*** for a given ***η*** is:

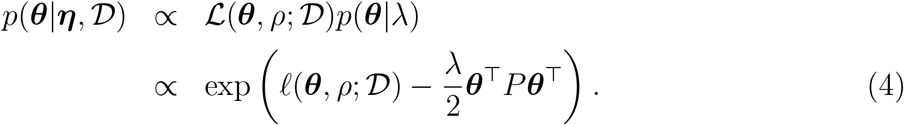

The gradient and Hessian of the log-likelihood (3) can be used to compute the gradient and Hessian of the (log-)conditional posterior (4), namely:

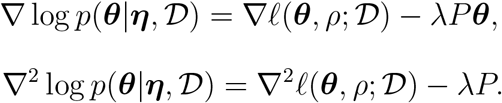

The above two equations will be used in a Newton-Raphson algorithm to obtain the Laplace approximation to the conditional posterior of ***θ***:

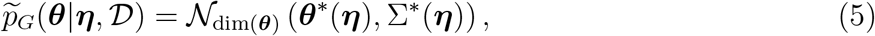

where ***θ****\**(***η***) and Σ*\**(***η***) is the mode and variance-covariance respectively after convergence of the Newton-Raphson algorithm. The latter two quantities are functions of the hyperparameter vector ***η***. An intuitive choice for ***η*** is to fix it at its *maximum a posteriori* (MAP). This is the option retained here, although it is also possible to work with a grid-based approach (Rue et al., 2009; Gressani and Lambert, 2021).

### 2.3 Hyperparameter optimization

The hyperparameter vector ***η*** = (*λ, ρ*)^*⊤*^ will be calibrated by posterior optimization. Following Tierney and Kadane (1986) and Rue et al. (2009), the hyperparameter vector can be approximated as follows:

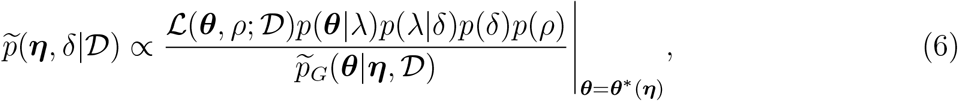

Approximation (6) can be written extensively as follows:

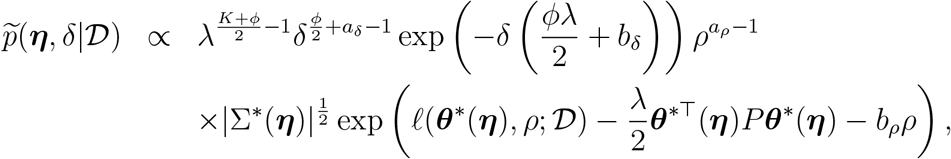

where the *K/*2 power of *λ* comes from the determinant 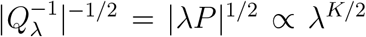. As 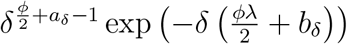 is the kernel of a Gamma distribution for the dispersion parameter *δ*, the following integral can be analytically solved:

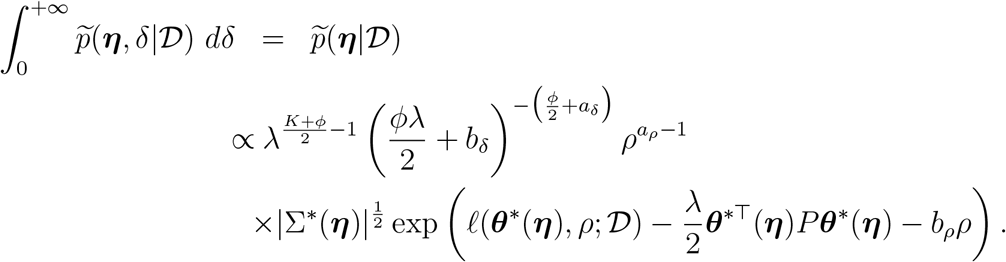

Using the transformation of variables (ensuring numerical stability during optimization) *w* = log(*ρ*), *v* = log(*λ*), one can show that 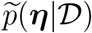 can be written as follows after using the multivariate transformation method:

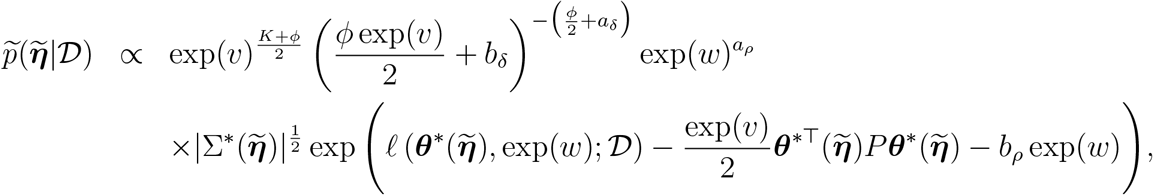

where 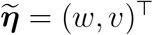. The approximated log-posterior becomes:

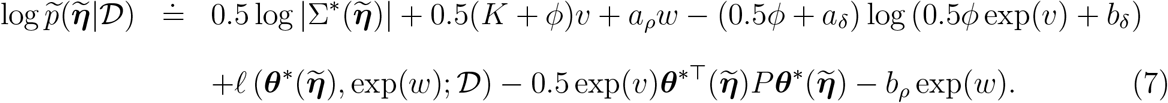

Equation (7) is numerically optimized and yields 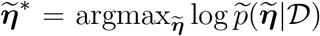. Plugging the latter vector into the Laplace approximation (5), one obtains the estimate 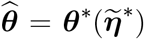 of the spline vector. The latter can be seen as a *maximum a posteriori* (MAP) estimate of ***θ***. Thus, the approximated posterior of the spline vector is:

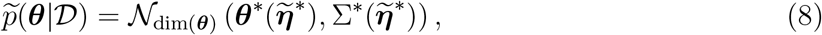

and can be used to construct credible intervals for functions that indirectly depend on ***θ***, such as *R*(*t*) as shown in the following section.

## 3 Estimation of *R*(*t*) with LPSMAP

### 3.1 The renewal equation “plug-in” estimate

In this section, we show how the Negative Binomial model for smoothing incidence counts can be used to estimate *R*(*t*) through the renewal equation. Let ***φ*** = {*φ*_1_, …, *φ*_*k*_} be a known *k*-dimensional vector representing the serial interval distribution. The renewal equation describes the link between the number of new cases at day *t* and past infected cases (up to time point *t*−1) weighted by the serial interval distribution. Mathematically:

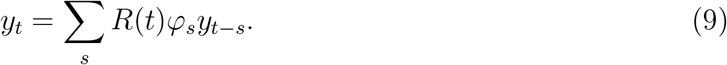

Rearranging (9) and taking the length *k* of the serial interval into account, we obtain an equation with the instantaneous reproduction number on the left-hand side:

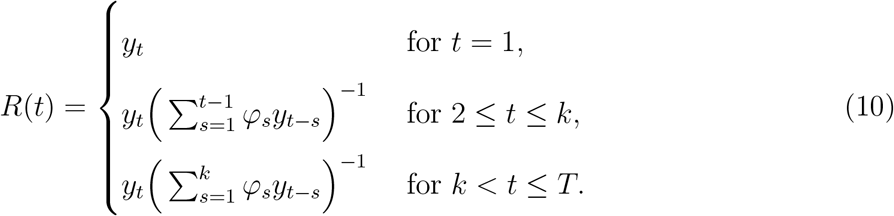

A “plug-in” estimator of *R*(*t*) at any *t* ∈ 𝒯 is obtained by replacing the number of new cases *y*_*t*_ by the estimated mean number of cases 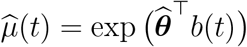, yielding:

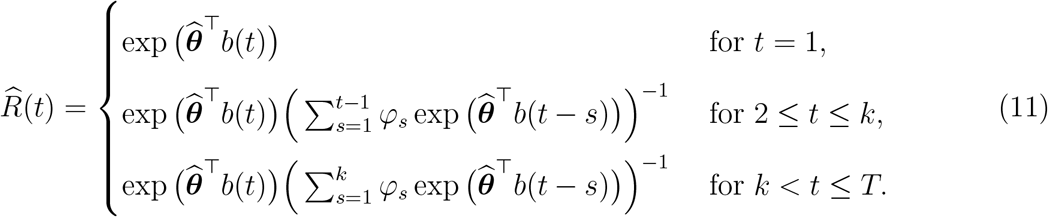

Using the indicator function 𝕀(·), i.e. 𝕀(*A*) = 1 if condition *A* is true and 𝕀(*A*) = 0 otherwise, the above “plug-in” estimate of *R*(*t*) can be written in a single line:

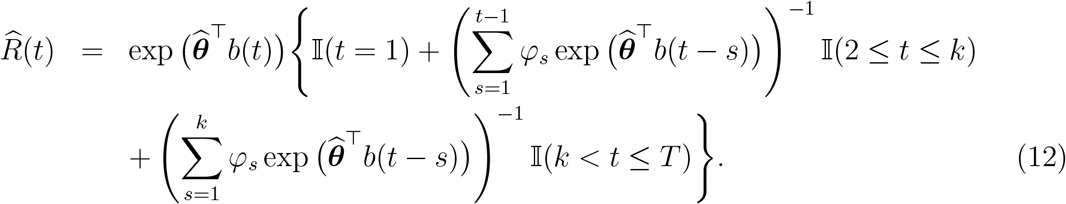

### 3.2 Credible intervals for *R*(*t*)

Replacing the number of new cases *y*_*t*_ by the (theoretical) mean number of cases *μ*(*t*) = exp(***θ***^*⊤*^*b*(*t*)) in (10) and using the compact notation, one has:

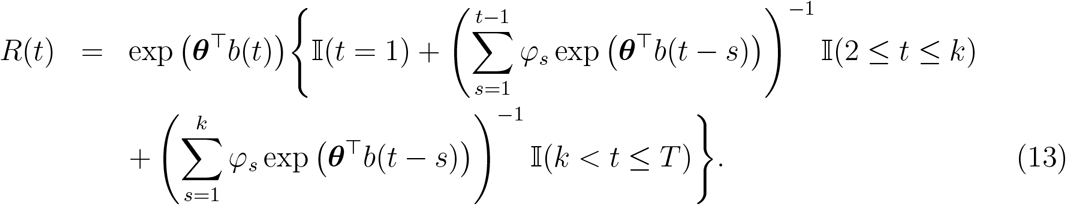

Let us denote the log of the instantaneous reproduction number in (13) as:

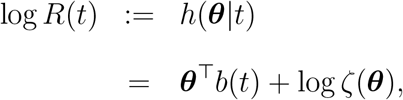

with

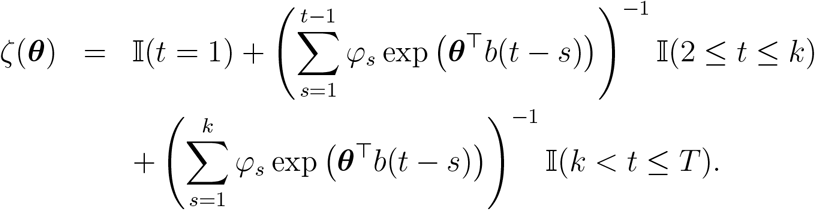

Note that *h*(***θ***|*t*) is seen here as a function of the spline vector ***θ*** for a given time point *t* ∈ 𝒯. A (1 *− α*) *×* 100% approximate credible interval for *R*(*t*) is obtained via a “delta” method. Consider a first-order Taylor expansion of *h*(***θ***|*t*) around 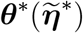 (henceforth ***θ****\** for the sake of a light notation), the mean of the Laplace approximated posterior of the spline vector in (8):

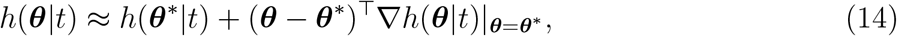

where the *k*th entry of the gradient vector ∇*h*(***θ***|*t*) = (*∂h*(***θ***|*t*)*/∂θ*_1_, …, *∂h*(***θ***|*t*)*/∂θ*_*K*_)^⊤^ is:

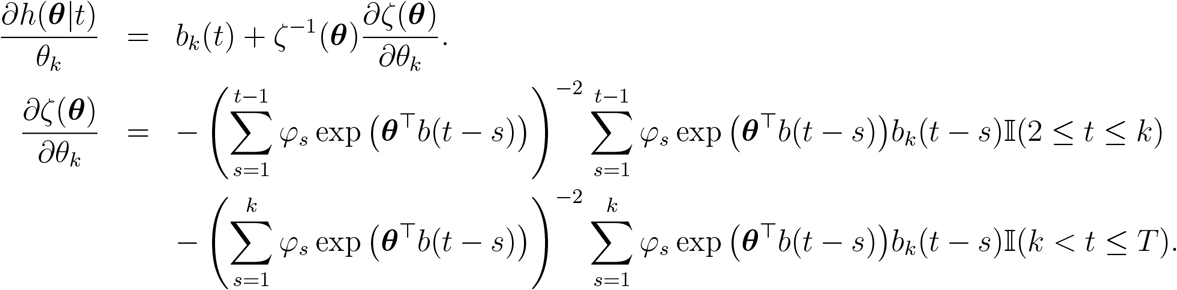

It follows that for *k* = 1, …, *K* we have:

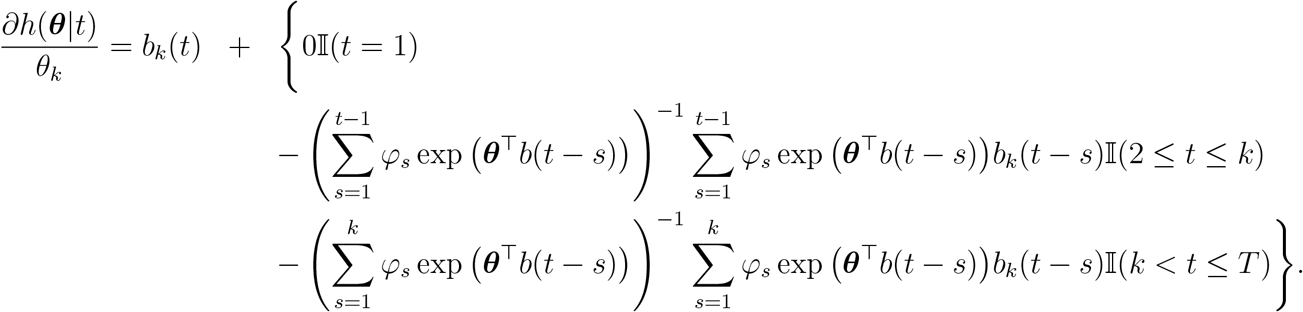

The Taylor expansion in (14) is a linear combination of the vector ***θ*** that is *a posterior* (approximately) Gaussian due to the Laplace approximation. As the family of Gaussian distributions is closed under linear combinations, it follows that *h*(***θ***|*t*) (and hence log *R*(*t*)) is *a posteriori* also (approximately) Gaussian with mean 𝔼(*h*(***θ***|*t*)) *≈ h*(***θ****\**|*t*) and variance-covariance matrix V(*h*(***θ***|*t*)) ≈ ∇^⊤^*h*(***θ***|*t*)|_***θ***=***θ****\**_Σ*\**∇*h*(***θ***|*t*)|_***θ***=***θ****\**_, where 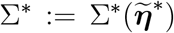 is the covariance matrix of the Laplace approximation (8). This suggests to write:

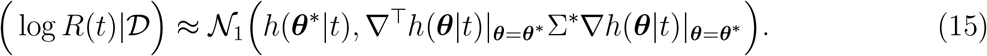

Thus, from (15) a quantile-based (1 *− α*) *×* 100% approximate credible interval for *R*(*t*) is:

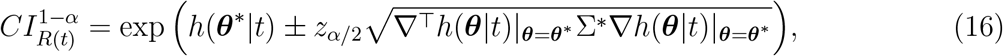

where *z*_*α/*2_ is the *α/*2-upper quantile of a standard normal variate.

## 4 Estimation of *R*(*t*) with LPSMALA

In Bayesian statistics, posterior distributions obtained with Bayes’ theorem often entail a high degree of analytical complexity and to a certain extent a flavor of incompleteness in the sense that the posterior is typically only known up to a normalizing constant. Markov chain Monte Carlo (MCMC) methods are simulation techniques for generating samples from a (possibly unnormalized) target distribution (Chib and Greenberg, 1996). One of the most popular MCMC methods together with the Gibbs sampler (Geman and Geman, 1984) is the Metropolis-Hastings (MH) algorithm originally proposed by Metropolis et al. (1953) and later generalized by Hastings (1970). In this section, we propose to implement a modified version of the Metropolis-adjusted Langevin algorithm (MALA) (Roberts and Tweedie, 1996) within the EpiLPS framework. The major advantage of MALA as compared to MH algorithms is that the proposal distribution is based upon a discretized approximation of the Langevin diffusion that uses the gradient of the target posterior distribution. These “smarter” proposals make use of additional information about the target density so that algorithms based on Langevin dynamics can converge at sub-geometric rates and tend to be more efficient than naive random-walk Metropolis algorithms (Roberts and Rosenthal, 1998, 2001).

This motivates our choice for embedding a MALA algorithm in EpiLPS as an efficient way of obtaining MCMC samples for inference on the instantaneous reproduction number *R*(*t*) via the renewal equation. The end-user will thus have a fully flexible choice regarding the underlying approach for estimating *R*(*t*) either via Laplacian-P-splines where the uncertainty surrounding the parameter *λ* responsible for smoothing is ignored and fixed at its “maximum a posteriori” (LPSMAP); or via a modified MALA algorithm where the uncertainty surrounding the penalty (and overdispersion) parameter is fully taken into account (LPSMALA). The approach permits to obtain samples from the joint posterior of the spline vector and the penalty and overdispersion parameters. The latter can then be injected in functionals of the spline vector and hence obtain smooth estimates of the epidemic curve as well as the instantaneous reproduction number. Another advantage is that highest posterior density intervals can be easily calculated from LPSMALA.

### 4.1 Conditional posteriors for a “Metropolis-within-Gibbs”

#### 4.1.1 Joint posterior of (*ζ, λ*)

Let ***ζ*** = (***θ***^⊤^, *ρ*)^⊤^ be the (*K* + 1)-dimensional vector gathering the B-spline coefficients ***θ*** and the overdispersion parameter *ρ*. Using Bayes’ theorem, the joint posterior distribution for ***ζ***, *λ* and *δ* is:

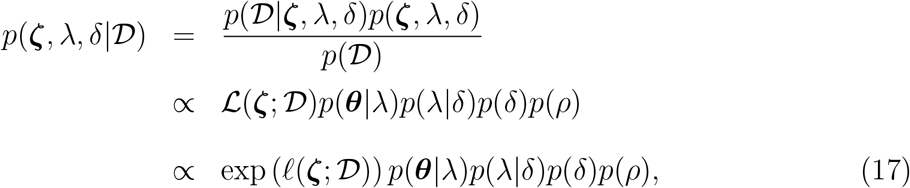

The analytical formulas of the chosen priors are:

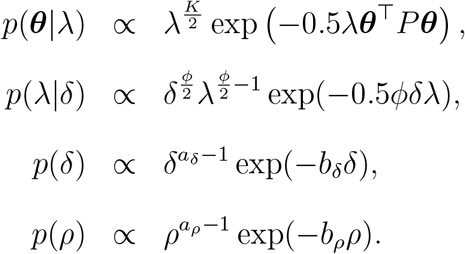

Injecting the above priors into (17) yields:

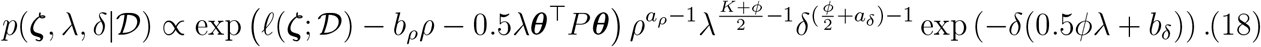

#### 4.2.1 Condtional posteriors of *ζ, λ* and *δ*

The following conditional posterior distributions can be directly obtained from (18):

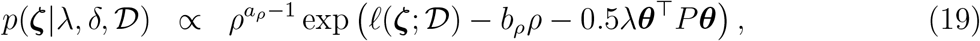

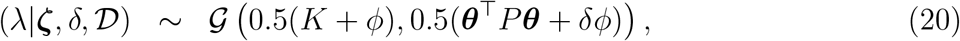

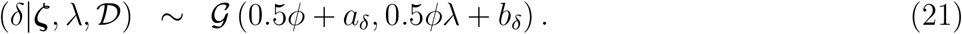

### 4.2 Sampling from the joint posterior *p*(*ζ, λ, δ*|*D*)

As the full conditionals *p*(***ζ***|*λ, δ*, 𝒟), *p*(*λ*|***ζ***, *δ*, 𝒟) and *p*(*δ*|***ζ***, *λ*, 𝒟) are available, we follow a “Metropolis-within-Gibbs” strategy to sample the joint posterior *p*(***ζ***, *λ, δ*|𝒟). In particular, the hyperparameters *λ* and *δ* will be sampled in a Gibbs step, while ***ζ*** will be sampled using a modified Langevin-Hastings algorithm. This approach is presented in Lambert and Eilers (2009) in the context of Bayesian density estimation (see also Lambert and Eilers (2005) for the use of MALA in a proportional hazards model). We adapt the algorithm of the latter reference to our EpiLPS methodology by incorporating the added value of our Laplacian-P-splines method. In particular, the variance-covariance matrix in the Langevin diffusion will be replaced by the variance-covariance matrix of LPSMAP. The correlation structure borrowed from LPSMAP improves convergence and chain mixing.

### 4.3 The modified Metropolis-adjusted Langevin algorithm

In what follows, we prefer to work under the log(·) parameterization for *ρ*, i.e. *w* = log(*ρ*) and denote by 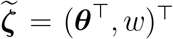, the (*K* + 1)-dimensional vector of B-spline amplitudes and overdispersion *w*. Under this parameterization, the conditional posterior of 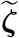 given *λ* and *δ* can be obtained from (19) by using the transformation method of random variables:

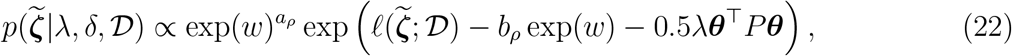

with the following log-likelihood under the reparameterization:

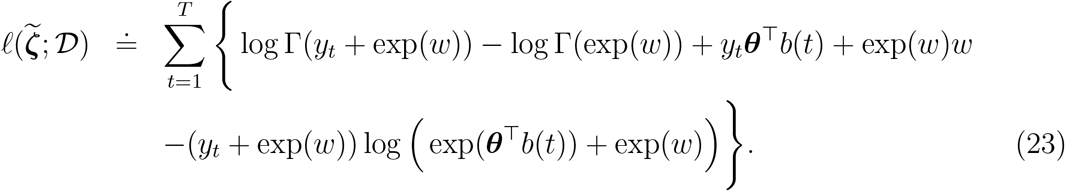

Let us denote by 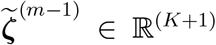 the state of the chain at iteration (*m −* 1). In the Langevin-Hastings algorithms, the proposal for the vector 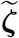 at iteration *m* is a draw from the following multivariate Gaussian distribution:

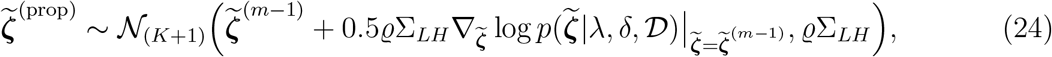

where *ϱ >* 0 is a tuning parameter that has to be carefully chosen in order to reach a desired acceptance rate and Σ_*LH*_ is the following block-diagonal variance-covariance matrix:

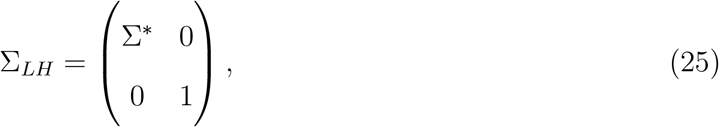

where Σ*\** is the *K*-dimensional covariance matrix obtained with LPSMAP. The gradient of log 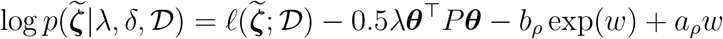 can be decomposed as follows:

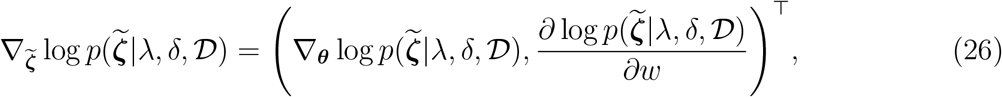

and is analytically available (see Appendix A for more details). All the quantities related to the Langevin-Hastings proposal have been analytically derived, so that the draw in (24) can be obtained (for a given value of *λ* and *δ*). As in a classic MH algorithm, the next step consists in computing the acceptance probability:

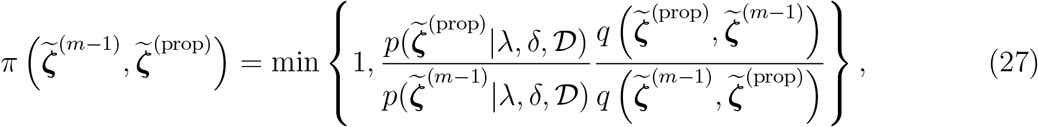

where *q*(*·, ·*) denotes the (Gaussian) proposal distribution and *p*(·|*λ, δ, D*) the target (conditional) posterior distribution. Finally, we generate a uniform random variable *u ∼ 𝒰* (0, 1) and accept the proposed vector 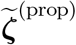 if 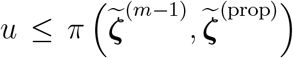 and reject it otherwise. While iterating through the Metropolis-within-Gibbs algorithm, the tuning parameter *ϱ* is automatically adapted to reach the optimal acceptance rate of 0.57 (Haario et al., 2001; Atchadé and Rosenthal, 2005; Roberts and Rosenthal, 1998). The pseudo-code below summarizes the LPSMALA algorithm.

#### LPSMALA algorithm to sample the posterior *p*(*θ, ρ, λ, δ*|𝒟)

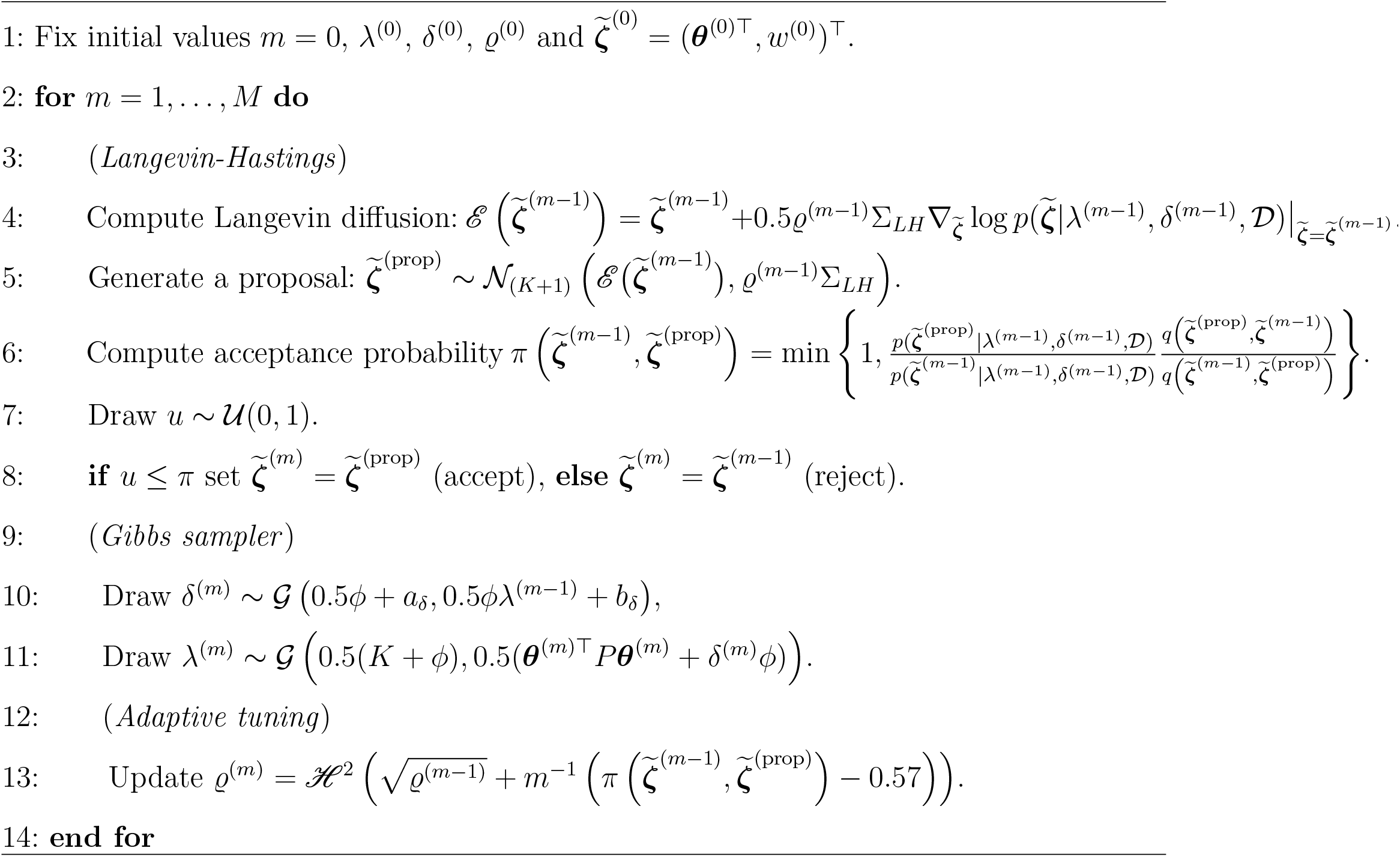

The adaptive tuning part (on line 13) involves the step function *ℋ* (*z*) = *ϵ*𝕀(*z < ϵ*) + *z*𝕀(*ϵ ≤ z ≤ 𝒜*) + *𝒜* 𝕀(*z > 𝒜*), with *ϵ* = 10^−4^ and *𝒜* = 10^4^, see Lambert and Eilers (2009) for details. Finally, the ratio 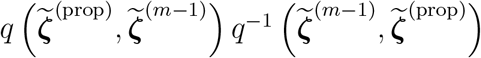 entering the computation of the acceptance probability (line 6) is derived in Appendix B.

### 4.4 Posterior inference

Provided the LPSMALA algorithm (cf. Section 4.3) is iterated long enough, say after 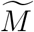 iterations, MCMC theory certifies that 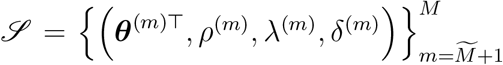 can be viewed as random draws from the target posterior distribution *p*(***θ***, *ρ, λ, δ*|𝒟). Note that a convenient starting point for the initial values of the parameters might be to fix them at their LPSMAP estimate. Given the sample *ℒ*, inference on quantities that are functions of ***θ*** becomes straightforward in the sense that point estimates and credibility envelopes can be easily obtained. A point estimate for the mean number of incidence counts at time *t* is taken to be the posterior mean (after discarding the burn-in phase):

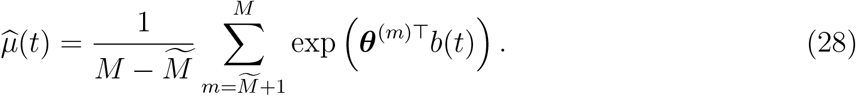

Note also that *ℒ* can be used to compute highest posterior density intervals of *μ*(*t*) at any point *t*. Using the renewal equation and the MCMC sample, one can apply the “plug-in” estimate of Section 3.1 and recover the following estimate of the instantaneous reproduction number at any given point *t*:

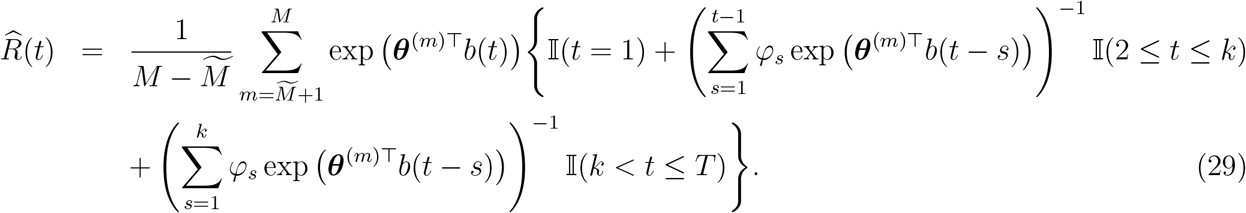

Also, using *ℒ*, one can compute a highest posterior density interval of *R*(*t*) at any point *t*.

## 5 Numerical study

In this section, we implement a numerical study with four underlying epidemic scenarios to assess the accuracy with which EpiLPS is able to “track” the target reproduction number over time. EpiLPS results are compared with the estimate R() routine of the EpiEstim package using two sliding window options (the default weekly window of 7 days and a shorter window of 3 days). In each scenario, we simulate incidence data for an epidemic lasting 50 days according to a Poisson data generating process with mean counts governed by the renewal equation (see Eq.10). In total, *S* = 200 epidemics are simulated for each scenario and a cubic B-spline basis of size *K* = 20 with a second order penalty is used for inference in EpiLPS. In Scenario 1, a constant instantaneous reproduction number at *R*(*t*) = 2.5 is considered. In Scenario 2, an intervention strategy is replicated, so that *R*(*t*) = 2.5 and a sudden drop to *R*(*t*) = 0.7 occurs at day *t* = 20. The latter scenario allows to check whether EpiLPS is able to quickly react to such a brutal change in *R*(*t*). Scenario 3 implements a more wiggly structure for *R*(*t*) and finally, Scenario 4 considers the case of a vanishing epidemic with a monotonic decreasing reproduction number. Table 1 summarizes the target *R*(*t*) functions and the serial interval distribution for the considered scenarios. Table 2 reports for each scenario the average Bias, MSE, coverage of 90% and 95% credible envelopes and credible interval width for the *R*(*t*) estimator with EpiLPS (LPSMAP and LSPMALA) and EpiEstim. The average is computed over days *t* = 8, …, 50. For LPSMALA, a chain of length 5, 000 and a warm up period of 2, 000 is specified.

**Table 1:** Different scenarios for the simulation of epidemic data based on a Poisson data generating process for the incidence time series over *T* = 50 days.

| Scenario | Target $R(t)$ | Serial interval distribution |
| --- | --- | --- |
| 1 | $R(t) = 2.5$ | $\varphi = (0.05, 0.05, 0.1, 0.2, 0.2, 0.1, 0.1, 0.1, 0.1)$ |
| 2 | $R(t) = 2.5 \mathbb{I}(t < 20) + 0.7 \mathbb{I}(t \geq 20)$ | $\varphi = (0.15, 0.25, 0.35, 0.13, 0.12)$ |
| 3 | $R(t) = 0.4 + \exp(\cos(t/5))$ | $\varphi = (0.13, 0.22, 0.18, 0.15, 0.07, 0.13, 0.12)$ |
| 4 | $R(t) = \exp(\cos(t/15))$ | $\varphi = (0.32, 0.24, 0.15, 0.09, 0.2)$ |

**Table 2:** Numerical results for EpiLPS and EpiEstim under different simulation scenarios. The Bias, MSE, 90% and 95% coverage of credible intervals and 95% CI width are averaged over days *t* = 8, …, 50.

| Scenario | Method | Bias | MSE | CP<sub>90%</sub> | CP<sub>95%</sub> | CI<sub>95%</sub> width |
| --- | --- | --- | --- | --- | --- | --- |
| Scenario 1<br>“Constant $R(t)$” | LPSMAP | -0.026 | 0.018 | 98.756 | 99.407 | 0.946 |
|  | LPSMALA | -0.026 | 0.020 | 93.535 | 95.977 | 0.351 |
|  | EpiEstim (7d window) | -0.003 | 0.016 | 90.291 | 95.221 | 0.335 |
|  | EpiEstim (3d window) | 0.003 | 0.036 | 90.444 | 95.567 | 0.499 |
| Scenario 2<br>“Step $R(t)$” | LPSMAP | -0.008 | 0.038 | 89.337 | 90.651 | 0.395 |
|  | LPSMALA | -0.005 | 0.030 | 90.384 | 90.814 | 0.328 |
|  | EpiEstim (7d window) | 0.085 | 0.071 | 76.733 | 81.302 | 0.123 |
|  | EpiEstim (3d window) | 0.030 | 0.036 | 86.000 | 90.691 | 0.253 |
| Scenario 3<br>“Wave $R(t)$” | LPSMAP | -0.007 | 0.009 | 97.372 | 98.895 | 0.530 |
|  | LPSMALA | 0.000 | 0.009 | 92.058 | 95.919 | 0.329 |
|  | EpiEstim (7d window) | 0.051 | 0.145 | 19.895 | 24.395 | 0.321 |
|  | EpiEstim (3d window) | 0.039 | 0.050 | 60.957 | 67.638 | 0.532 |
| Scenario 4<br>“Decaying $R(t)$” | LPSMAP | 0.000 | 0.001 | 98.477 | 99.465 | 0.257 |
|  | LPSMALA | -0.001 | 0.001 | 90.767 | 95.419 | 0.112 |
|  | EpiEstim (7d window) | 0.125 | 0.021 | 17.988 | 20.616 | 0.097 |
|  | EpiEstim (3d window) | 0.046 | 0.010 | 46.436 | 51.819 | 0.214 |

For the first scenario, EpiEstim exhibits slightly lower bias than EpiLPS and the MSE is more or less of the same magnitude for both approaches. A general pattern that is apparent from Table 2 is that EpiLPS tends to overcover with LPSMAP (except for Scenario 2), while LPSMALA is closer to the nominal value in all scenarios. This phenomenon can be attributed to the following two main sources of approximations inherent to LPSMAP: (1) the Laplace approximation to the conditional posterior of the B-spline amplitudes and (2) the *maximum a posteriori* estimate of the hyperparameter vector that ignores the uncertainty related to the estimation of the roughness penalty parameter. In Scenario 2, EpiLPS has smaller bias than EpiEstim and the latter approach also suffers from mild undercoverage with a weekly sliding window. In Scenarios 3 and 4, where the target *R*(*t*) curve is less linear than in the previous scenarios, a more pronounced difference between EpiEstim and EpiLPS appears. EpiLPS shows better performance, especially in terms of coverage of credible intervals where EpiEstim exhibits severe undercoverage. Even when decreasing the sliding window to 3 days, the coverage probability is far from the nominal value.

Figures 1-4 show the simulated incidence data together with the estimated trajectories of *R*(*t*) with EpiLPS (using LPSMAP) and EpiEstim respectively. Under linear configurations of *R*(*t*), the two competing approaches have similar behavior, while for curved *R*(*t*) functions, EpiEstim estimates (with a weekly sliding window) are shifted as compared to the target. With EpiEstim, the magnitude of this shift can be controlled by decreasing the sliding window, yet the optimal *a priori* choice of a smoothing window is far from being an easy task in practice and thus alleviating the window size assumption (as in EpiLPS) can be of interest.

**Figure 1:**
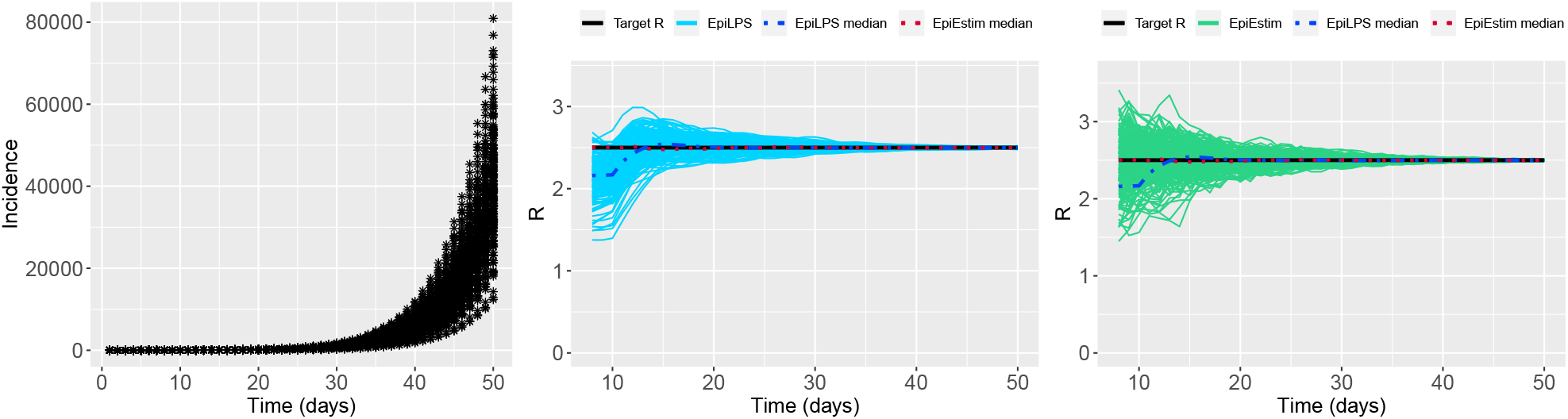
(Left) Simulated incidence data for Scenario 1. (Center) Estimated trajectories of *R*(*t*) for each generated dataset with EpiLPS (LPSMAP). (Right) Estimated trajectories of *R*(*t*) for each generated dataset with EpiEstim and a weekly sliding window. The pointwise median estimate of *R*(*t*) for EpiLPS (dashed) and EpiEstim (dotted) is also shown.

**Figure 2:**
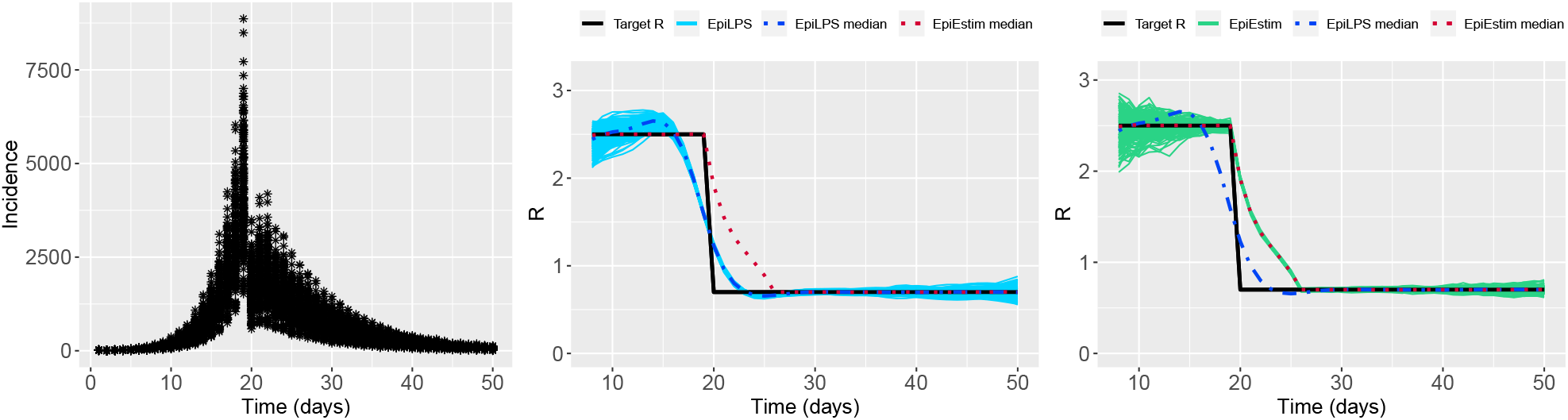
(Left) Simulated incidence data for Scenario 2. (Center) Estimated trajectories of *R*(*t*) for each generated dataset with EpiLPS (LPSMAP). (Right) Estimated trajectories of *R*(*t*) for each generated dataset with EpiEstim and a weekly sliding window. The pointwise median estimate of *R*(*t*) for EpiLPS (dashed) and EpiEstim (dotted) is also shown.

**Figure 3:**
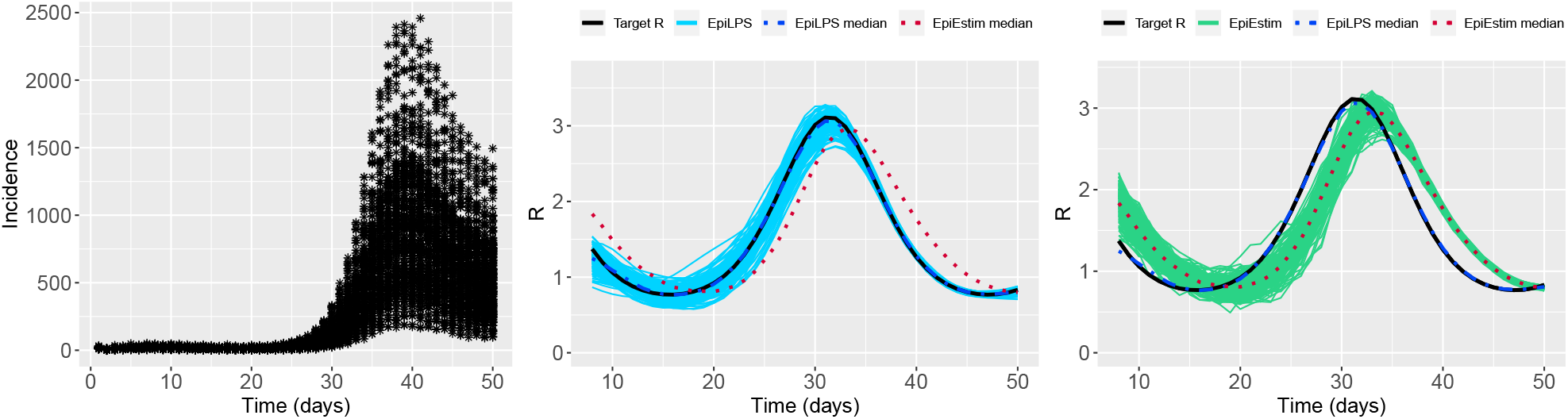
(Left) Simulated incidence data for Scenario 3. (Center) Estimated trajectories of *R*(*t*) for each generated dataset with EpiLPS (LPSMAP). (Right) Estimated trajectories of *R*(*t*) for each generated dataset with EpiEstim and a weekly sliding window. The pointwise median estimate of *R*(*t*) for EpiLPS (dashed) and EpiEstim (dotted) is also shown.

**Figure 4:**
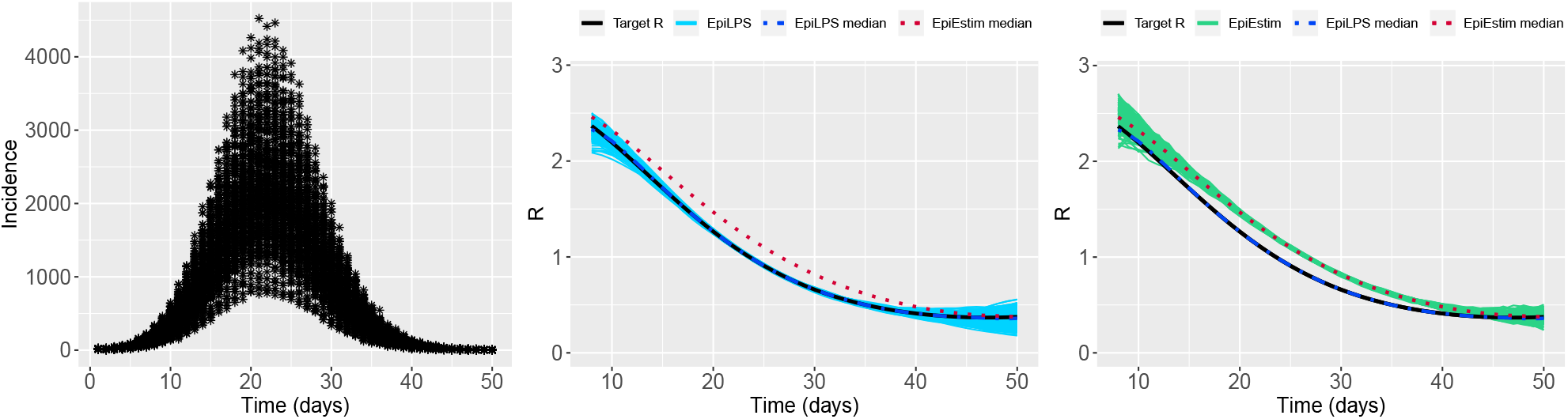
(Left) Simulated incidence data for Scenario 4. (Center) Estimated trajectories of *R*(*t*) for each generated dataset with EpiLPS (LPSMAP). (Right) Estimated trajectories of *R*(*t*) for each generated dataset with EpiEstim and a weekly sliding window. The pointwise median estimate of *R*(*t*) for EpiLPS (dashed) and EpiEstim (dotted) is also shown.

## 6 Application to observed case counts in infectious disease epidemics

### 6.1 Epidemics of SARS-CoV-1 and influenza A H1N1

In this section, the LPSMALA algorithm is applied on two historical outbreak datasets presented in Cori et al. (2013). In particular, we consider the 2003 SARS outbreak in Hong Kong and the 2009 pandemic influenza in a school in Pennsylvania (USA). We use *K* = 40 B-splines with a second-order penalty and the serial interval distributions provided in the EpiEstim package Cori (2021). The LPSMALA algorithm is implemented with a chain of length 25,000. Acceptance rates for the generated chains are close to the optimal value of 57% and the posterior samples have converged according to the Geweke (1992) diagostic test (at the 1% level of significance). Figure 5 shows the smoothed epidemic curves and the estimated *R*(*t*) for the two outbreaks. Results for the SARS data show that the reproduction number reaches a first peak during the third week where 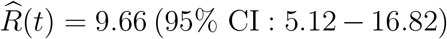 and a second more moderate peak around week 6 with 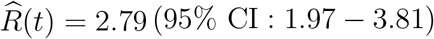. After day *t* = 43, the epidemic is under control and *R*(*t*) smoothly decays below 1. For the pandemic influenza in Pennsylvania, in the end of the second week *R*(*t*) is around 2.04 (95% CI : 1.24 *−* 3.07). During the middle of the third week, the situation is less severe and *R*(*t*) points below 1. As noted in Cori et al. (2013), a few cases appeared in the last days of the epidemic generating an upward trend in *R*(*t*) estimates.

**Figure 5:**
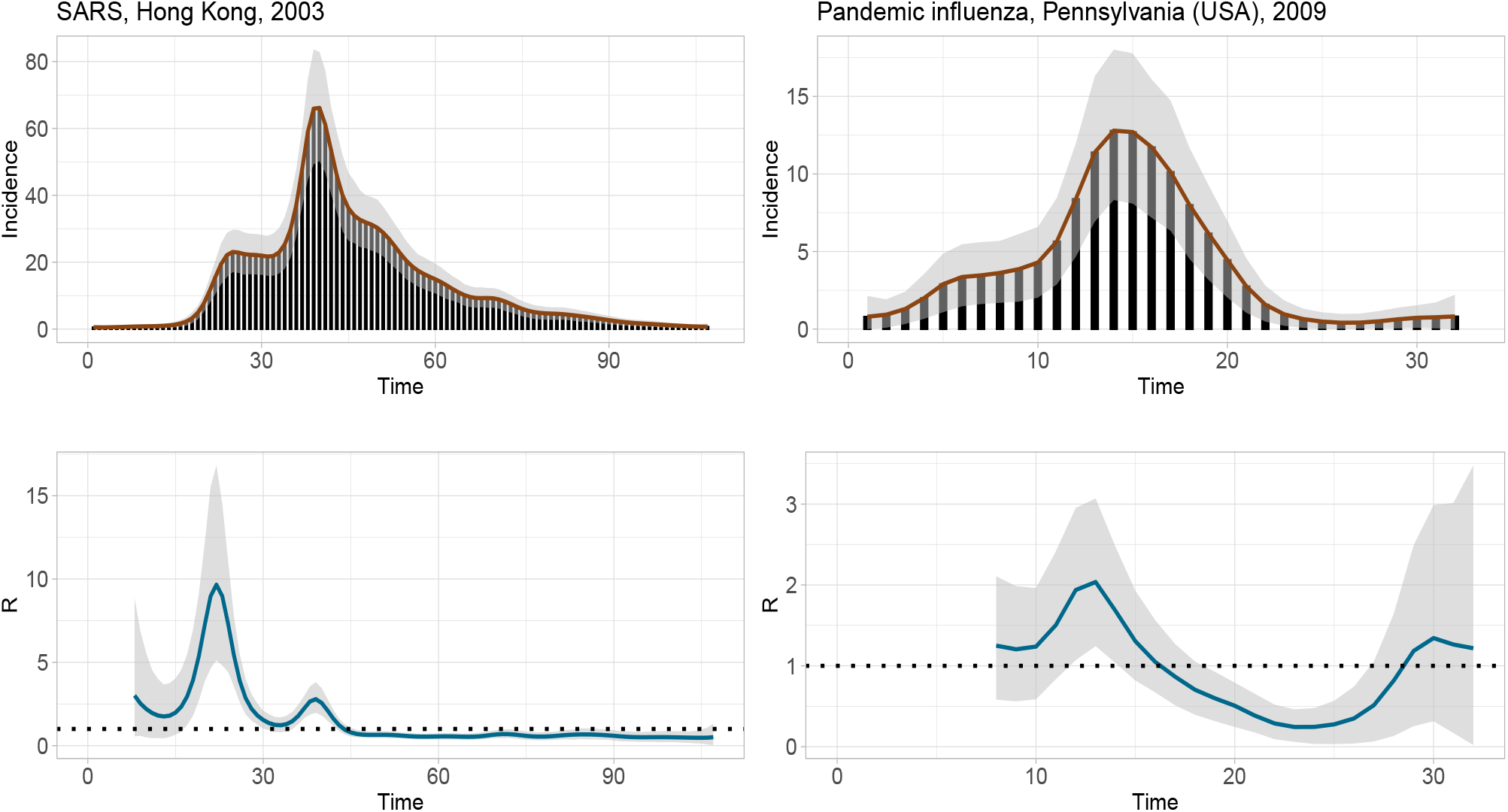
(Left column) EpiLPS fit for the epidemic curve (top) and the instantaneous reproduction number *R*(*t*) (bottom) of the SARS outbreak in Hong Kong, 2003. (Right column) EpiLPS fit for the epidemic curve (top) and the instantaneous reproduction number *R*(*t*) (bottom) of the pademic influenza in Pennsylvania, 2009.

### 6.2 Application on the SARS-CoV-2 pandemic

The EpiLPS methodology is illustrated on the SARS-CoV-2 pandemic using publicly available data from the Covid-19 Data Hub (Guidotti and Ardia, 2020) and its associated COVID19 package on CRAN (https://cran.r-project.org/package=COVID19). Countrylevel data on hospitalizations for Belgium, Denmark, Portugal and France from April, 5th 2020 to October 31st, 2021 is used with a uniform serial interval distribution over five days, i.e. ***φ*** = (0.2, 0.2, 0.2, 0.2, 0.2). In Figure 6, the estimated reproduction number over the considered period is shown for the four countries. Results are obtained with the LPSMAP algorithm using *K* = 30 B-splines and a second order penalty. From a computational perspective, it takes less than 3 seconds to fit the EpiLPS model for the four countries with LPSMAP. The fitted reproduction numbers reflect the different waves of the COVID-19 pandemic and the recent rise in infections in the beginning of September 2021.

**Figure 6:**
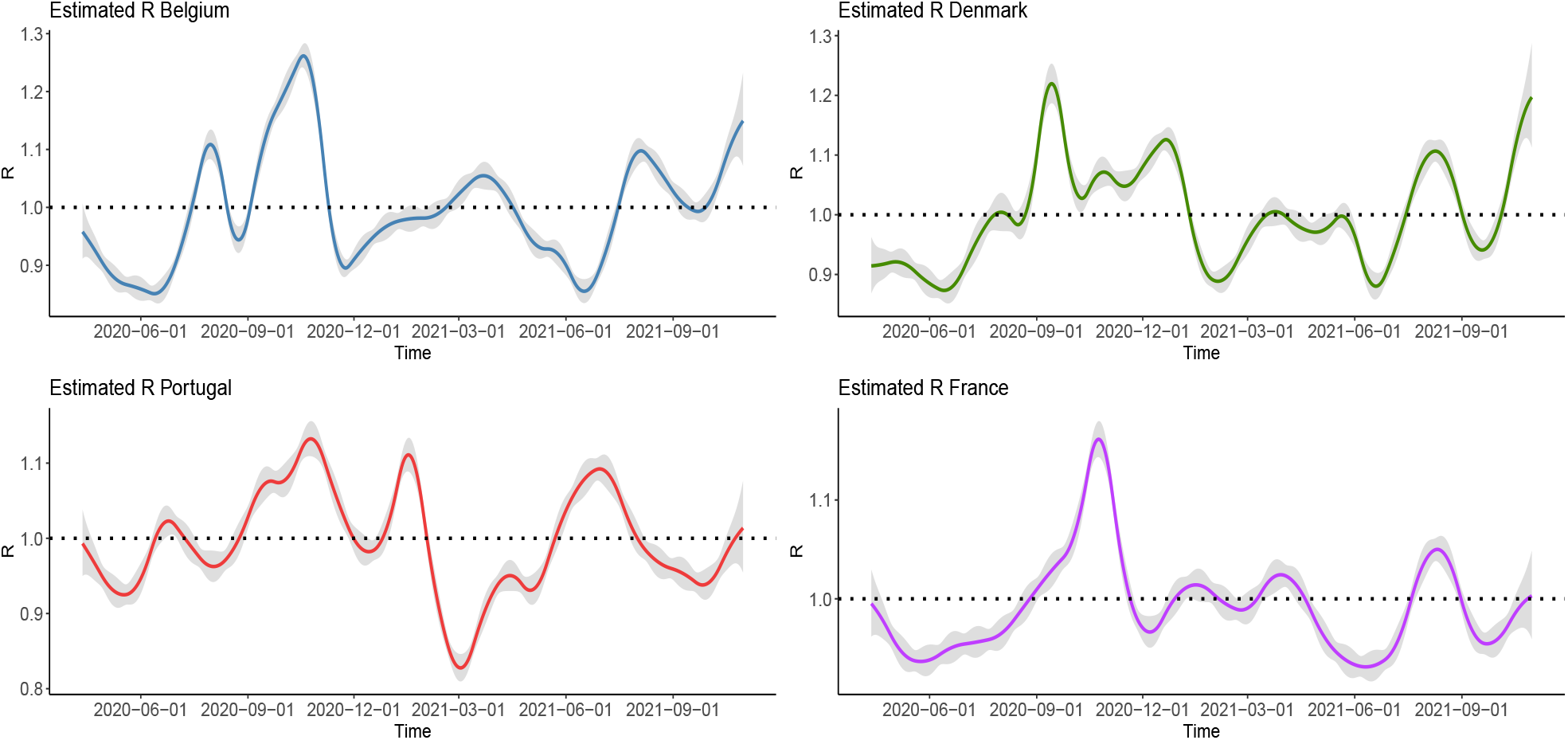
Estimated reproduction number from 2020-04-05 to 2021-10-31 for Beglium, Denmark, Portugal and France using a five days serial interval and LPSMAP with *K* = 30 B-splines and a second order penalty. The gray shaded surface corresponds to the 95% (approximate) credible interval.

## 7 Conclusion

EpiLPS (an acronym for **Epi**demiological modelling with **L**aplacian-**P**-**S**plines) is a fast and flexible tool for near real-time estimation of the instantaneous reproduction number *R*(*t*) during epidemic outbreaks. The tool is flexible in the sense that (penalized) spline based approximations provide smoothed estimates of *R*(*t*) with little computational effort and without the constraint of imposing any sliding window assumption that could potentially affect the timing and accuracy of the estimator. Moreover, the end user has the choice between a fully sampling-free approach (LPSMAP) or an efficient MCMC-based approach with Langevin diffusions (LPSMALA) for inference. The available EpiLPS package (https://github.com/oswaldogressani) allows public health policy makers to analyze incoming data faster than existing methods relying on classic MCMC samplers, thus allowing them to be better informed when taking decisions on control measures for the ongoing SARS-CoV-2 epidemic. Simulation studies in this manuscript provide encouraging results and support EpiLPS as being a robust tool capable of a precise tracking of *R*(*t*) over time. The available EpiLPS software package is also straightforward to use with simple function calls that are specifically interesting for routine usage.

The EpiLPS project opens up several future research directions. A possible extension would be to formulate the EpiLPS model within a zero-inflated Poisson framework to cope with incindence time series characterized by an excess of zero counts. Another interesting extension would be to adapt the model to account for regional variation and imported cases.

## Data Availability

Simulation results and real data applications in this manuscript can be fully reproduced
with the EpiLPS package available here https://github.com/oswaldogressani.

## Software

Simulation results and real data applications in this manuscript can be fully reproduced with the EpiLPS package available here https://github.com/oswaldogressani.

## Acknowledgments

This project is funded by the European Union’s Research and Innovation Action under the H2020 work programme, EpiPose (grant number 101003688).

## Appendix A

In this appendix, we derive the analytical expressions of the gradient:

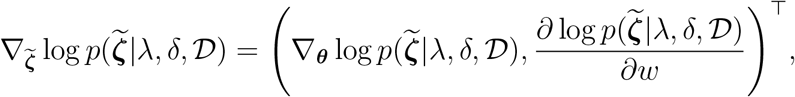

where the target function is 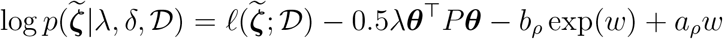. Let us first concentrate on the partial derivatives with respect to the spline components:

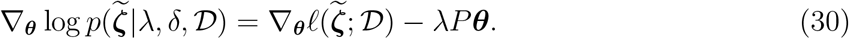

As already shown in Section 2.2, the gradient for the log-likelihood 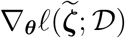 is:

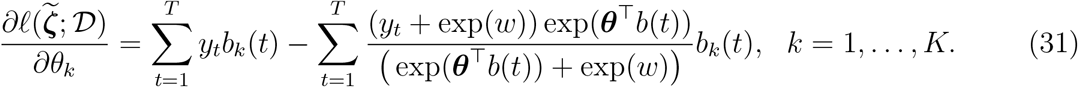

The last term to be computed to recover the full gradient is:

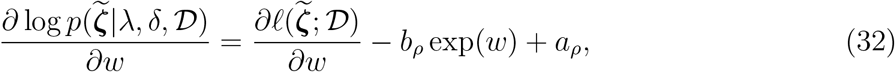

Where

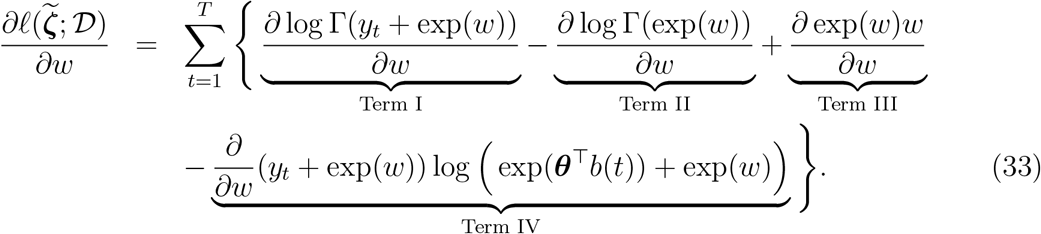

For Term I, using the chain rule, one recovers:

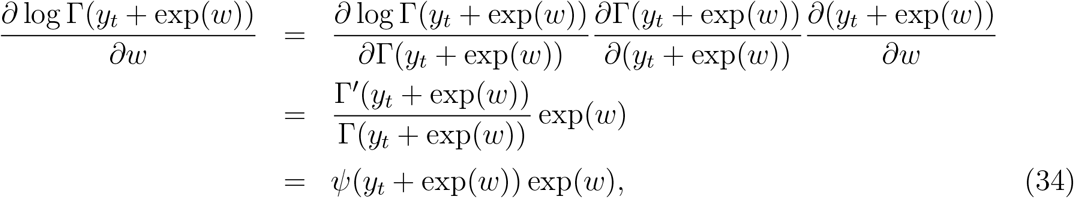

where *ψ*(·) is the digamma function. Using the same chain rule argument, one can easily show that for Term II:

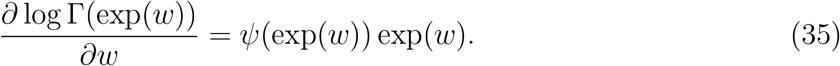

Term III is also trivial:

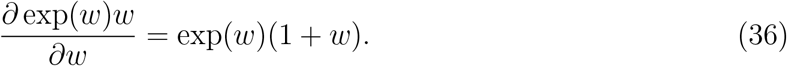

Term IV is as follows:

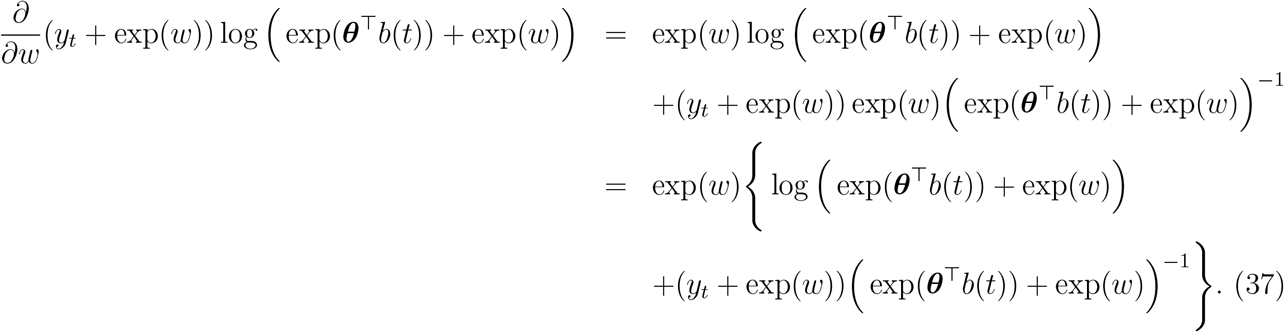

Gathering all the above intermediate results, we obtain the following derivative for (32):

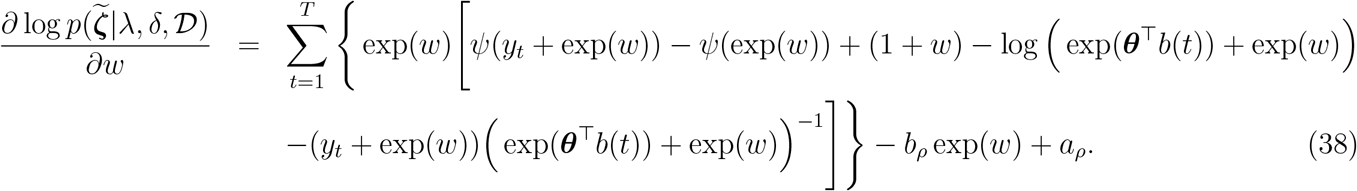

## Appendix B

To determine the analytical form of the ratio of proposal distributions in the Metropolis-within-Gibbs algorithm, let us use the compact notation 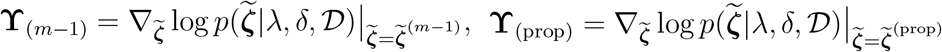, and define the difference 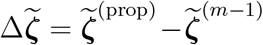, so that:

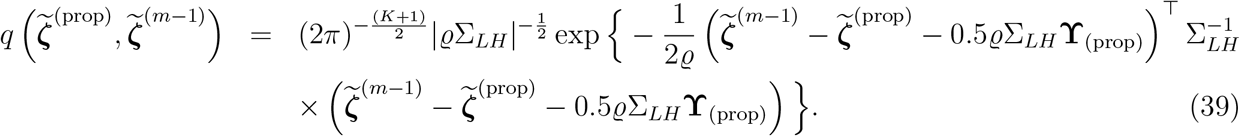

The kernel of (39) is thus:

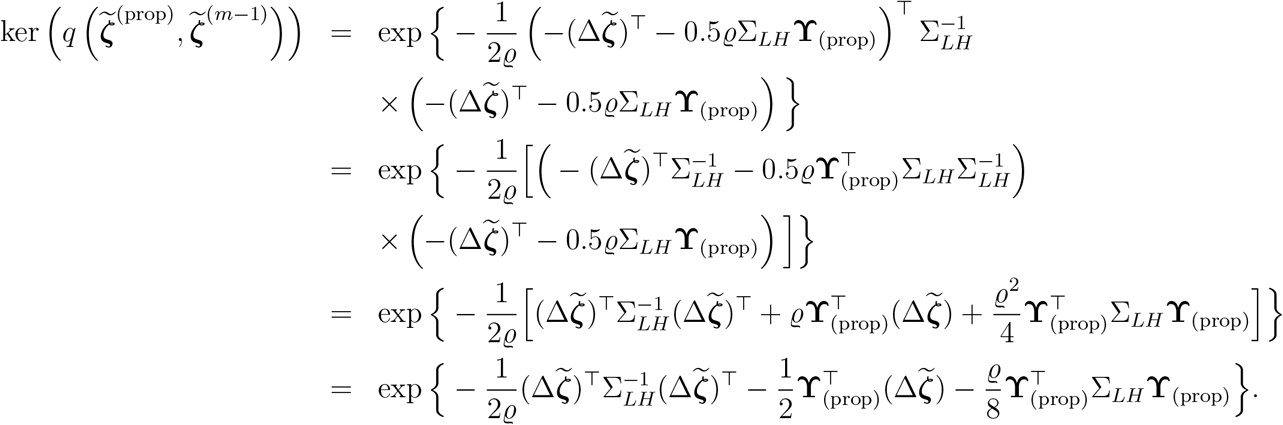

Using a similar argument, one can show that the kernel of 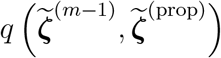 is:

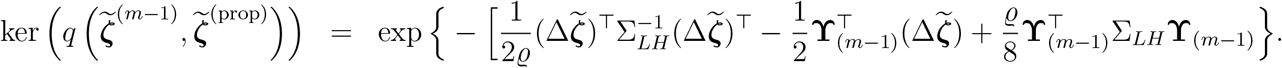

This can be used to compute the ratio:

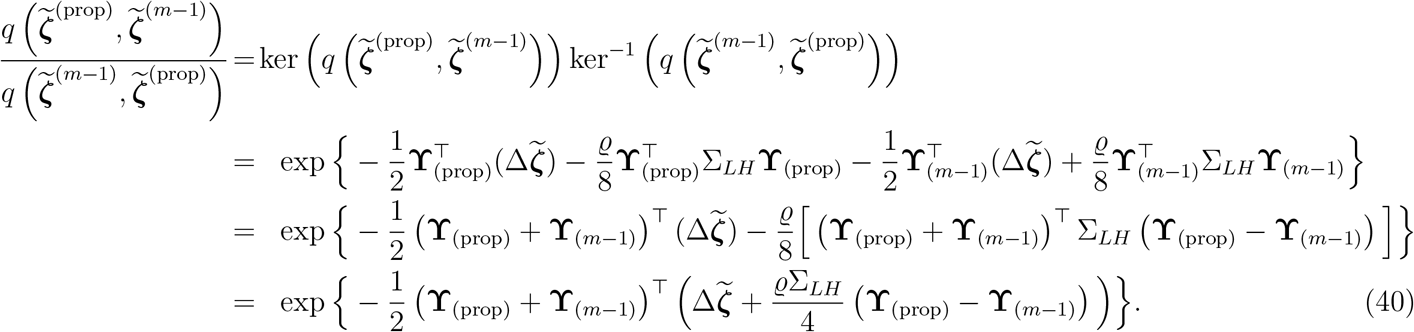

